# Impact of school-based malaria screening and treatment on *P. falciparum* infection and anemia prevalence in two transmission settings in Malawi

**DOI:** 10.1101/2021.04.22.21253119

**Authors:** Lauren M. Cohee, Ingrid Peterson, Jenna E. Coalson, Clarissa Valim, Moses Chilombe, Andrew Ngwira, Andy Bauleni, Sarah Schaffer-DeRoo, Karl B. Seydel, Mark L. Wilson, Terrie E. Taylor, Don P. Mathanga, Miriam K. Laufer

**Affiliations:** Center for Vaccine Development and Global Health, University of Maryland School of Medicine, Baltimore, MD, USA; Department of Epidemiology, School of Public Health, University of Michigan, Ann Arbor, MI, USA; Eck Institute for Global Health, University of Notre Dame, Notre Dame, IN, USA; Department of Global Health, Boston University School of Public Health, Boston, MA, USA; Malaria Alert Center, University of Malawi College of Medicine, Blantyre, Malawi; University of Maryland School of Medicine, Baltimore, MD, USA; Children’s National Medical Center, Washington, DC, USA; College of Osteopathic Medicine, Michigan State University, East Lansing, MI, USA

**Keywords:** chemoprevention, anemia, schoolchildren, adolescent, intervention

## Abstract

**Background:** In areas highly endemic for malaria, *Plasmodium falciparum* (*Pf*) infection prevalence peaks in school-age children, adversely affecting their health and education. School-based intermittent preventive treatment reduces this burden, however concerns about cost and widespread use of antimalarial drugs have limited enthusiasm for this approach. School-based screening-and-treatment is an attractive alternative. We conducted a school-based cohort study to evaluate the impact of screening-and-treatment on the prevalence of *Pf* infection and anemia in two different transmission settings.

**Methods:** We screened 704 students in four Malawian primary schools for *Pf* infection using rapid diagnostic tests (RDTs). Those testing positive were treated with artemether-lumefantrine. Outcomes were *Pf* infections detected by microscopy or PCR and anemia after six weeks.

**Results:** Prevalence of infection by RDT at screening was 37% (range among schools 9-64%). We detected a significant reduction after six weeks in infections by microscopy (adjusted relative reduction (aRR) 47.1%, p<0.0001) and PCR (aRR 23.1%, p<0.0001), but no reduction in anemia. In low, seasonal prevalence areas, sub-patent infections at screening led to persistent infection, but not disease, during follow-up. In high transmission settings, new infections frequently occurred within six weeks after treatment.

**Conclusions:** School-based screening-and-treatment reduced *Pf* infection but not anemia. This approach could be enhanced in low transmission settings by using more sensitive screening tests and in high transmission settings by repeating the intervention or using longer acting drugs.

**Summary:** Malaria screening-and-treatment reduced *P. falciparum* infections but not anemia. Because rapid diagnostic tests failed to detect low-density infections, screening tests with higher sensitivity may be needed in low transmission areas where low-density infections make-up a larger proportion of infections.

## Background

Despite global declines in morbidity and mortality over the last two decades, malaria remains a significant public health burden in most of sub-Saharan Africa.^1^ Malaria surveillance and research has largely focused on children less than five-years old and pregnant women, the groups at highest risk of adverse outcomes. Expansion of surveillance to other age-groups has revealed an under-appreciated burden of *Plasmodium falciparum (Pf)* infection in school-age children.^2^ While severe disease is less likely in school-age children when compared with younger children, infection in school-age children is associated with anemia^3–9^, increased school absenteeism^10–12^, and lower performance on cognitive testing^3,12–15^.

School-based preventive treatment of malaria is a promising strategy to reduce *Pf* infection, clinical malaria, and anemia in school-age children.^16^ Preventive treatment includes both “intermittent preventive treatment”, in which all students are treated regardless of infection status, and “screen-and-treat”, in which students are screened for infection with a point of care test and treated if positive. To our knowledge, the only clinical trial evaluating the screen-and-treat approach in schoolchildren demonstrated no decrease in prevalence of infection or anemia.^17^ One explanation for the lack of efficacy in this large, cluster-randomized trial is the use of a relatively short-acting treatment, artemether-lumefantrine (AL) (duration of prevention 13.8 days^18^) and students were screened once per school term, allowing substantial time for re-infection. Furthermore, the rapid diagnostic tests (RDTs) used for screening are not sensitive in detecting low-density infections, which are common in school-age children and also comprise a larger proportion of infections in low prevalence settings, such as coastal Kenya where this trial took place.^19^ Nevertheless, a screen-and-treat approach has multiple advantages, including limiting treatment to test-positive children, which decreases the cost and adverse effects of medication and the selective pressure for drug resistance. Given these advantages, more evidence evaluating and optimizing the screen-and-treat approach will determine the potential role of screen-and-treat as a tool to maximize health.

We conducted a school-based, prospective, cohort study to evaluate the epidemiology of *Pf* infections in school-age children and determine the impact of the screen-and-treat approach on *Pf* infection and anemia prevalence among students in two different transmission settings. We hypothesized that RDTs frequently fail to detect low-parasite-density infections, and that these infections contribute considerably to the burden and health consequences of *Pf* infection in school-age children. We further hypothesized that school-age children are frequently reinfected following treatment, necessitating repeated screening and administration of antimalarial therapy to maintain the impact of the intervention. The relative importance of these potential barriers to the effectiveness of screen-and-treat interventions in schoolchildren may differ based on the transmission setting. Our ultimate goal is to inform the development of malaria control interventions to improve the health of school-age children.

## Methods

### Study sites

We conducted school-based cohort studies of students in four schools that were selected from malaria surveillance sites in southern Malawi.^20^ Two schools (in Maseya and Makhuwira) had high parasite prevalence in school-age children (>40% in the rainy and dry seasons of 2012-2014 and less than a twofold prevalence difference between seasons). The other two schools (in Bvumbwe and Ngowe) had lower, seasonally variable transmission (<15% prevalence in the rainy and dry seasons of 2012-2014 and at least a twofold difference between seasons). Cohorts were followed at the end of the rainy season (April-May) and during the dry season (September-October) of 2015, when *Pf* transmission is high and low, respectively. Routine malaria control measures in place during our study included long-lasting insecticide treated nets (LLINs) that were distributed nationally in 2012, and freely available malaria diagnosis (using RDTs) and treatment (with AL) in government-operated health facilities.

### Study design

Prior to initiating the study, sensitization meetings were held with community leaders, school leaders, and district and local government officials, who provided permission to conduct the study. Using a random number generator, 15 students per grade-level (grades 1-8), were invited to participate. Students were excluded if they did not have a parent/guardian available to provide consent, would not be attending the school for the duration of the study, or had a known allergy to AL. To evaluate the intervention separately in each season, students enrolled in the rainy season cohort were excluded from participating in the dry season cohort.

Visits were conducted at baseline (1-2 weeks after enrollment), and 1, 2 and 6 weeks later. At the baseline visit, a finger prick blood sample was obtained for point-of-care detection of infection using a histidine-rich protein 2-based RDT in accordance with manufacturer instructions (Paracheck Orchid Biomedical Systems, Goa, India or SD Bioline, Standard Diagnostics Inc., Suwon City, Republic of Korea), measurement of hemoglobin (Hemocue, Angelholm, Sweden), microscopy and molecular detection tests for *P. falciparum*. Students with infections detected by RDT were treated with AL (Novartis Pharma AG or Ajanta Pharma Ltd) using weight-based dosing. At each follow-up visit, another blood sample was obtained for microscopy and molecular detection of parasites, which was not conducted in real time, and a study team member conducted a one-on-one interview with each student about bed net use the night prior, current or recent illness, and use of antimalarial treatment. At the final visit, RDT and hemoglobin were repeated, and intercurrent fever or malaria treatment was identified by parent interview and review of portable medical records (“health passports”). Data was collected on android-based tablets using OpenDataKit and RedCap.^21^

### Sample size determination

Using surveillance data collected in the communities surrounding the intervention schools in the prior three years, we anticipated a 27% overall prevalence of *Pf* infection detected by PCR in school-age children. By sampling 320 students (10 students within each of 8 grades, in four schools) in each season, we would have 80% power to detect a 40% decrease in the overall prevalence of infection after the screen-and-treat intervention. To account for loss to follow-up, we sampled 15 students per grade in each school.

### Laboratory methods

Thick smear microscopy slides were stained with Giemsa stain and read by trained, blinded microscopists. Quality control was conducted using published procedures.^22^ Molecular detection of parasites was conducted using real-time PCR to detect *P. falciparum* lactate dehydrogenase (*Pf*LDH) DNA, as described previously.^23^ Plates were run in duplicate and the sample was considered positive if a positive result was detected in at least one run.

### Statistical analysis

*Pf* infection was analyzed as a binary variable. Infections detected by microscopy and PCR were analyzed separately. Sub-patent infections were defined as those that were PCR-positive but RDT-negative. Treatment groups were defined based on RDT result at baseline. RDT-positive students with negative PCR results were considered negative for active infection, but were included in the treatment group for analysis because they received AL in the intervention. Anemia was defined as hemoglobin less than 11.0 g/dL. Non-study administered malaria treatment at baseline or during follow-up was defined as any treatment with an effective antimalarial reported by the student, during the parent interview, or in the student’s health passport.

To analyze repeated measures of the following binary outcomes, we used log-binomial and logistic generalized estimating equation (GEE) models, with unstructured covariance and clustering within students nested in schools: PCR-detected *Pf* infection, microscopy-detected *Pf* infection, sub-patent infection, and fever in the past 48 hours. GEE models were used to estimate proportions and odds ratios. Logistic regression was used to model anemia at day 42, and a linear model was used to model hemoglobin at day 42; both models included adjustment for anemia status at baseline. All models were adjusted *a priori* for factors known to be associated with malaria risk: school (as a proxy for transmission intensity), age 5 to 9 years versus 10 to 15 years, and season of recruitment; other potential confounding variables were included in the model if they were significant at *P*<0.05. An interaction term for school*day was significant in models of PCR-detected *Pf* infection, microscopy-detected *Pf* infection and was included in these models. Relative difference in prevalence was calculated as the difference between prevalence at baseline and after six weeks divided by baseline prevalence. Analyses were conducted using SAS statistical software version 9.3 (SAS Institute, Cary, NC).

### Human subjects

Written informed consent and assent were obtained from guardians and children in accordance with Good Clinical Practice guidelines. Approval for this study was obtained from the University of Malawi College of Medicine Research and Ethics Committee and the Institutional Review Board of the University of Maryland, Baltimore.

## Results

A total of 786 students were enrolled: 405 in the rainy season cohort and 381 in the dry season cohort (Figure 1). Screening-and-treatment was conducted at baseline for 704 (90%) students, and complete follow-up was obtained for 616 (78%) students. Baseline characteristics, including sex, bed net use, anemia, recent fever and prevalence of *Pf* infection, varied significantly by school (Table 1). Students had increased odds of being RDT-positive and hence, receiving treatment, if they attended a higher prevalence school, participated in the rainy season cohort, were male, were anemic, or had recently taken an antimalarial medication (Table 2).

**Table 1.** Baseline characteristics of students enrolled in the study

|  | **Total**<br>**N=704** | **Bvumbwe**<br>**n=184** | **Ngowe**<br>**n=186** | **Maseya**<br>**n=185** | **Makhuwira**<br>**n=149** | **P<sup>a</sup>** |
| --- | --- | --- | --- | --- | --- | --- |
| Female, n(%) | 382(54.3) | 118(64.1) | 94(50.5) | 92(49.7) | 78(52.4) | 0.019 |
| Older school-age ($\geq 10$y), n(%) | 513(72.9) | 125(67.9) | 129(69.4) | 143(77.3) | 116(77.9) | 0.067 |
| Slept under bed net last night<sup>b</sup>, n(%) | 309(49.3) | 62(53.9) | 124(68.5) | 65(35.3) | 58(39.5) | <.0001 |
| Rainy season, n(%) | 364(51.7) | 99(53.8) | 100(53.8) | 97(52.4) | 68(45.6) | 0.411 |
| Anemic<sup>c</sup>, n(%) | 106(15.1) | 9(4.9) | 35(18.8) | 34(18.4) | 28(18.8) | 0.0002 |
| Hemoglobin in g/dL, mean (SD) | 12.3 (1.5) | 13.1 (1.3) | 12.0 (1.5) | 11.9 (1.3) | 11.9 (1.3) | <.0001 |
| Fever last 48 hours<sup>d</sup>, n(%) | 192(27.8) | 28(15.5) | 53(29.6) | 64(34.8) | 47(32.0) | 0.0002 |
| Recent malaria treatment<sup>e</sup>, n(%) | 121(17.4) | 25(13.7) | 33(18.2) | 35(19.0) | 28(19.1) | 0.479 |
| RDT Positive, n(%) | 259(36.8) | 16(8.7) | 56(30.1) | 91(49.2) | 96(64.4) | <.0001 |
| Sub-patent infection<sup>f</sup>, n(%) | 63(9.0) | 12(6.5) | 10(5.4) | 22(11.9) | 19(12.8) | 0.031 |

<sup>a</sup> P-value of differences across school<sup>b</sup> Net use data for Bvumbwe was only available in the dry season<sup>c</sup> Defined as hemoglobin <11.0 g/dL<sup>d</sup> Fever in the last 48 hours reported or measured temperature $\geq 37.5^{\circ}\text{C}$ <sup>e</sup> Recent treatment defined as student or parent reported receipt of an effective antimalarial drug in the two weeks preceding the interview<sup>f</sup> At baseline, 63 children had a negative RDT result but had a positive result for malaria PCR (n=61) or microscopy (n=2); higher proportion of *Pf* infections were sub-patent in Bvumbwe compared to the other three schools (42.8% [12/28] of infections were sub-patent vs. 17.3% [51/294], p=0.003)

**Table 2.** Associations with *P. falciparum* infections detected by RDT among enrolled students at baseline^a^

|  |  | n(%)<sup>b</sup> | Unadjusted<br>Prevalence Ratio | Adjusted Prevalence<br>Ratio |
| --- | --- | --- | --- | --- |
| School |  |  |  |  |
|  | Bvumbwe | 16(8.7) | 1 | 1 |
|  | Ngowe | 56(30.1) | 3.9(2.2-6.7) | 3.8(2.2, 6.6) |
|  | Maseya | 91(49.2) | 6.3(3.7-10.6) | 5.8(3.4, 9.8) |
|  | Makhuwira | 96(64.4) | 8.3(4.9-13.9) | 7.4(4.4, 12.4) |
| Season |  |  |  |  |
|  | Rainy | 159(43.7) | 1 | 1 |
|  | Dry | 100(29.4) | 0.7(0.6-0.8) | 0.7(0.6, 0.9) |
| Age (years) |  |  |  |  |
|  | 5 to 9 | 61(31.9) | 1 | 1 |
|  | 10 to 15 | 198(38.6) | 1.3(1.0-1.6) | 1.0(0.9, 1.3) |
| Sex |  |  |  |  |
|  | Male | 141(43.8) | 1 | 1 |
|  | Female | 118(30.9) | 0.7(0.6-0.9) | 0.9(0.7, 1.0) |
| Anemic<sup>c</sup> |  |  |  |  |
|  | No | 193(32.3) | 1 |  |
|  | Yes | 65(61.3) | 1.9(1.6-2.3) |  |
| Fever last 48 hours<sup>d</sup> |  |  |  |  |
|  | No | 170(34.1) | 1 |  |
|  | Yes | 84(43.8) | 1.3(1.0-1.6) |  |
| Recent medication<sup>e</sup> |  |  |  |  |
|  | No | 193(33.6) | 1 |  |
|  | Yes | 62(51.2) | 1.5(1.2-1.9) |  |
| Slept under bed net last night |  |  |  |  |
|  | No | 150(37.9) | 1 |  |
|  | Yes | 109(35.3) | 0.9(0.8-1.1) |  |

<sup>a</sup> Log-binomial model with school, season, age and sex retained in model *a priori*, all other variables retained if $p < .05$ . With the exception of incomplete data, univariate associations were assessed in 704 children, among whom 9 were missing data for recent medication, 13 were missing data for fever in last 48 hours, and 1 was missing data for anemia. The final model included 690 children.
<sup>b</sup> Number and percent of children with a positive RDT result within each strata; children with a positive RDT result at baseline were treated and children with a negative RDT result at baseline were not treated.
<sup>c</sup> Defined as hemoglobin less than 11.0 g/dL
<sup>d</sup> Fever in the last 48 hours reported or measured temperature $\geq 37.5^{\circ}\text{C}$
<sup>e</sup> Recent treatment defined as student or parent reported receipt of an effective antimalarial drug in the two weeks preceding the interview

**Figure 1:**
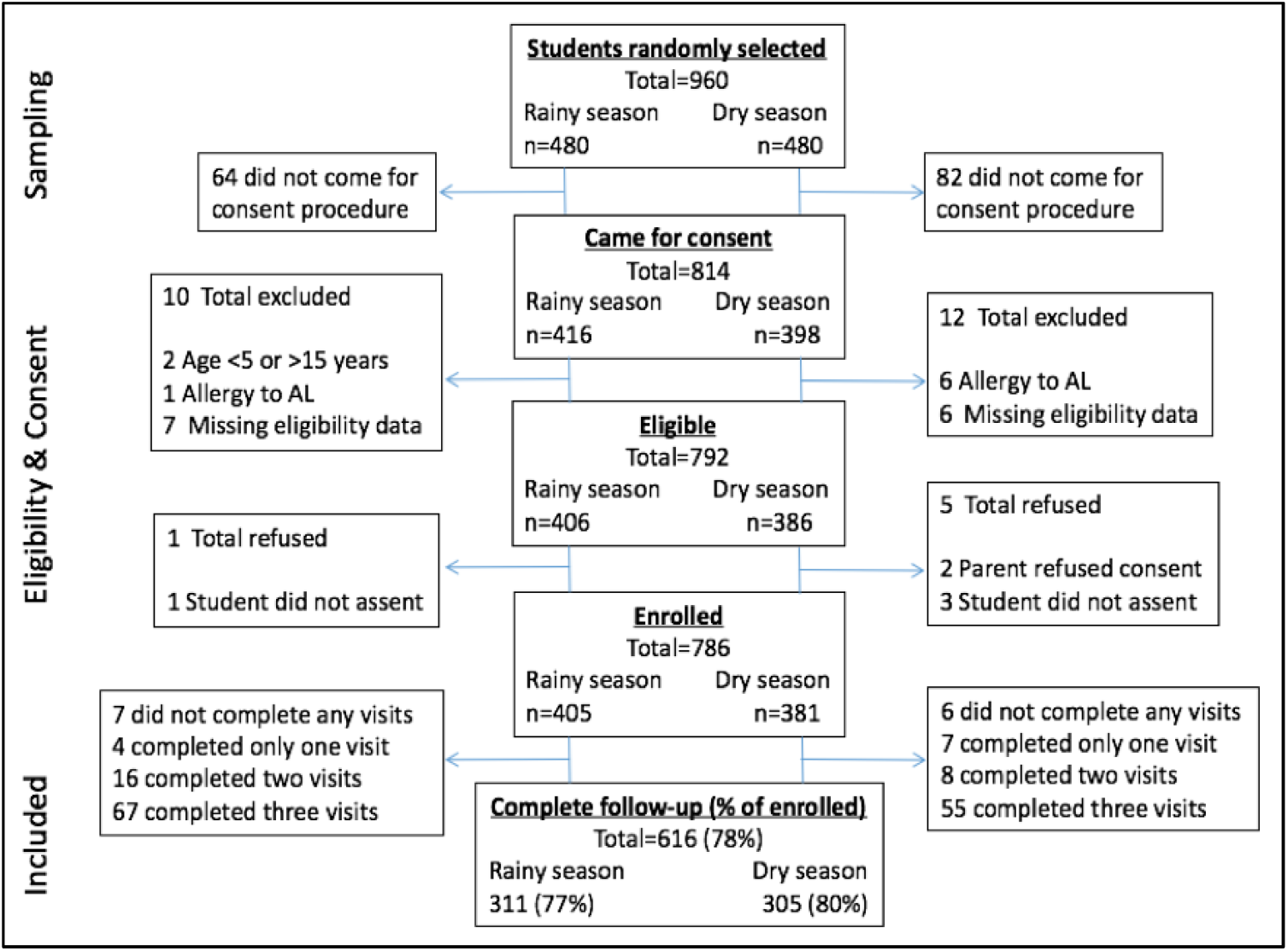
Enrollment and follow-up of participants in school-based cohorts

Sub-patent infections were common in the study population. Among the 259 students with infections detected by PCR or microscopy at baseline, 63 (24.3%) were negative by RDT. While these sub-patent infections were more likely to occur in the high transmission schools (Supplementary Table S1), they make up a larger proportion of all infections in Bvumbwe, the school with the lowest prevalence of infection [42.8% *vs*. 17.3%, (p=0.003) of infections were sub-patent Bvumbwe vs. the other three schools combined (Table 1)]. Compared to microscopy and PCR, respectively, RDTs had a sensitivity of 91.1% and 76.3% and specificity of 76.1% and 85.9%. We evaluated the impact of screening-and-treatment on *Pf* infections detected both by microscopy, to facilitate comparisons with prior studies and assess impact on infections more likely to be clinically relevant, and by PCR, to assess the impact including lower density infections.

### Impact of screening and treatment on prevalence of infection after six weeks

In total, 259 students (37%) were RDT-positive at baseline and were treated with AL. Six weeks after screening-and-treatment, we measured a 47.1% and 23.1% reduction in the prevalence of infections detected by microscopy and PCR, respectively (both p<0.001, Table 3). In analyses stratified by transmission setting, the prevalence of infections detected by microscopy decreased by 42.1% in high transmission schools and by 66.8% in low, seasonal transmission schools (p<0.001, p=0.003, respectively). For infections detected by PCR, we observed a 35.0% decrease in prevalence in the high transmission schools, but a non-statistically significant 13.3% increase in prevalence in the low, seasonal transmission schools (p<0.001, p=0.40, respectively). Results by school are shown in Figure S1.

**Table 3:**
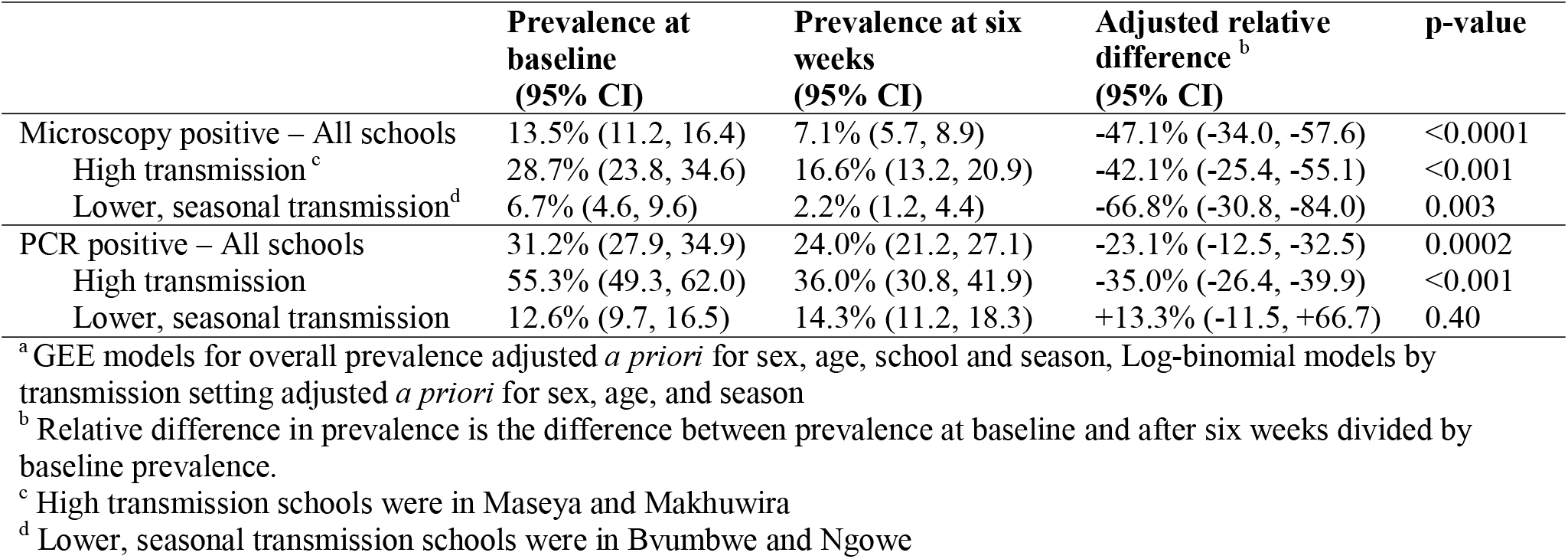
Adjusted prevalence and mean of infection outcomes at baseline and six weeks after the intervention^a^

### Patterns of infection and variation in the effect of screening-and-treatment by transmission setting

To further evaluate patterns of infection and the effect of screening-and-treatment in different transmission settings, we examined school-specific differences in the prevalence of infection during follow-up among treated and untreated students (Figure 2). In the higher transmission schools, RDTs detected most infections and treatment effectively cleared those infections at the one and two week follow-up visits. However, prevalence of infection among both treated and untreated students increased between two and six weeks following the intervention (Figure 3B and 3D).

**Figure 2:**
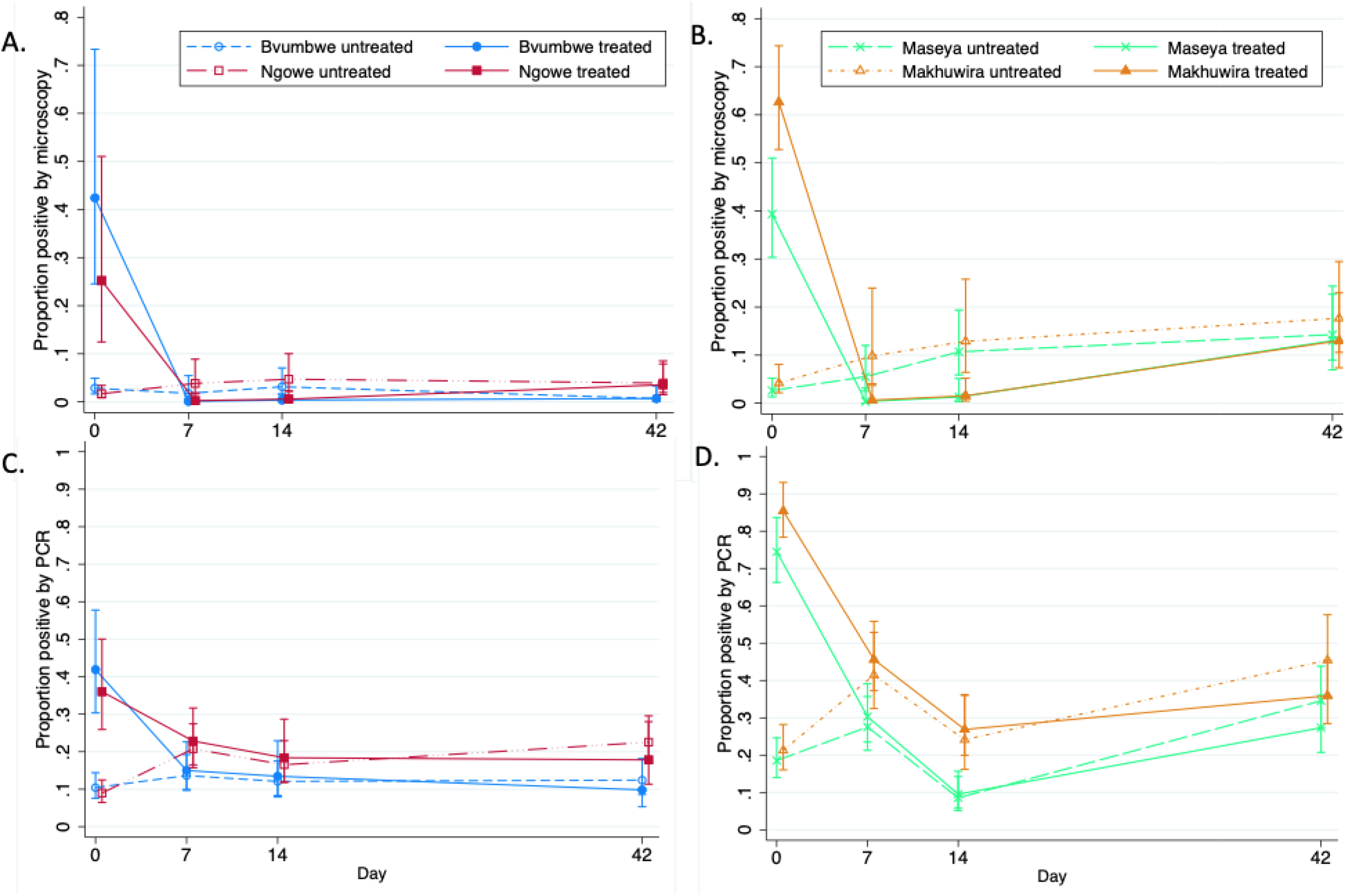
Proportion of *P. falciparum* infections stratified by school, treatment status, and detection method. Low, seasonal transmission schools (Bvumbwe-blue circles; Ngowe-red squared) and infections detected by microscopy (A) and PCR (C). High transmission schools (Maseya-green x’s; Makhuwira-orange triangles) and detection by microscopy (B) and PCR (D). Treated students-solid lines; Untreated students-hashed lines.

**Figure 3:**
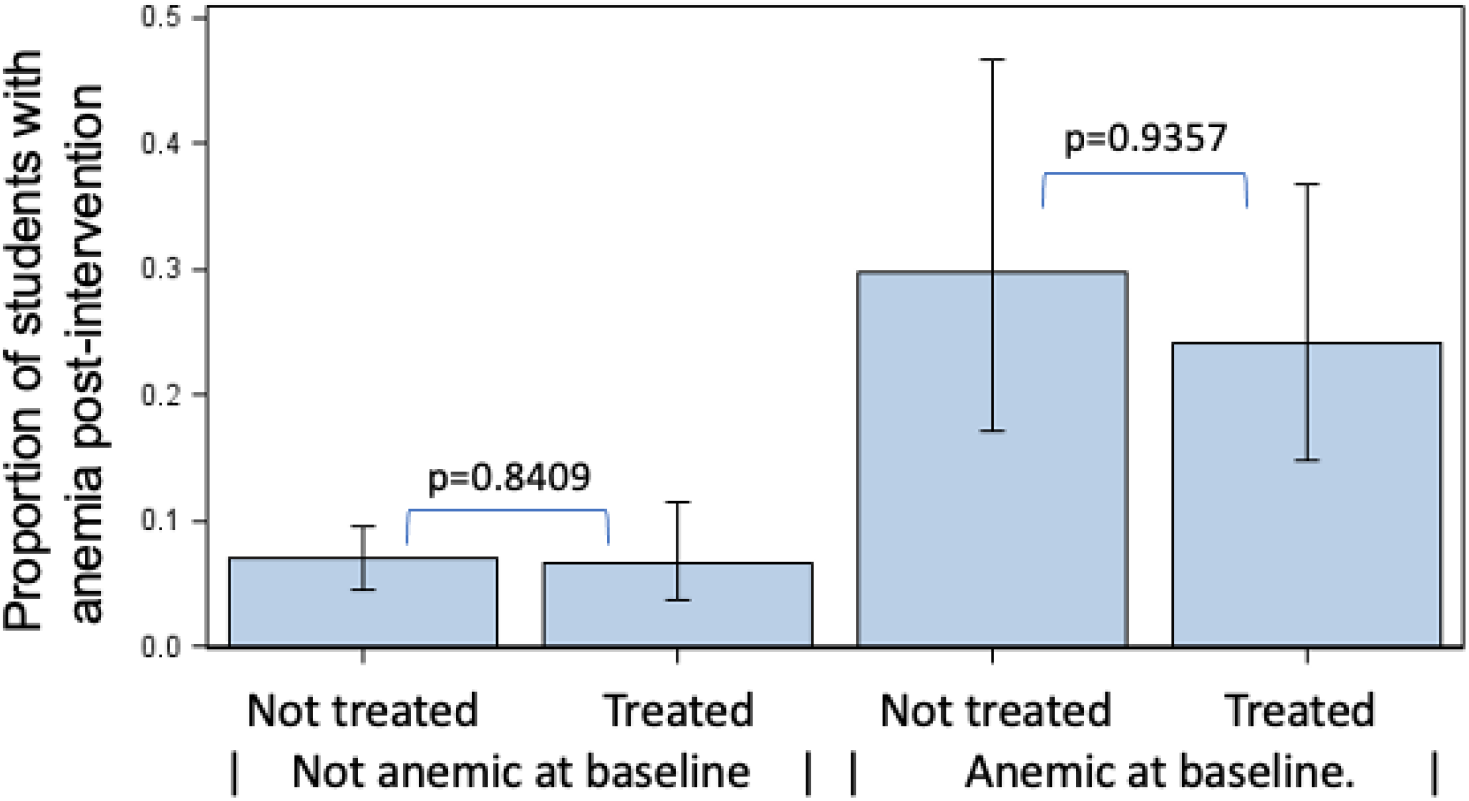
Proportion of students with anemia six weeks after the intervention by baseline anemia and treatment, adjusted for school, sex, season, and age. Unadjusted data in Supplement Figure S2.

In the schools with lower, seasonal transmission, 38% (22/58) of infections at baseline were sub-patent, and thus not detected by RDTs and therefore not treated. Treatment cleared infections detected by microscopy in the students who received treatment (Table 3, Figure 3A). However, there was no overall reduction in infections detected by PCR due to both persistence of infections detected by PCR in untreated students and either uncleared or new infections detected by PCR in the treated students (Table 3, Figure 3C).

### Impact of screening-and-treatment on anemia

At baseline, 15.1% (95% CI: 12.4%, 17.7%) of students were anemic and the mean hemoglobin was 12.3g/dL (SD: 1.5g/dL) (Table 1). Anemia at baseline was less common among students in Bvumbwe compared to those in other schools, and less common among girls than boys. However, baseline anemia was more common among students with a positive RDT and students with recent fever (Supplemental Table S2). In analysis accounting for baseline anemia and treatment status, risk of anemia six weeks after the intervention was not significantly associated with treatment status of the individual students (adjusted odds ratio (aOR): 1.14; 95% CI: 0.63, 2.05; *P*=0.5420, Figure 3).

### Clinical and parasitological outcomes of sub-patent infections

To evaluate the consequences of sub-patent infections, we compared clinical and parasitological outcomes among students with sub-patent infection to uninfected students, defined as students with negative RDT, microscopy, and PCR at screening. Sub-patent infections at screening were not associated with adverse clinical outcomes at the six week follow-up, including fever (aOR 1.05 [95%CI:0.64,1.72]), antimalarial treatment (aOR 0.97 [95%CI: 0.40, 2.32]), anemia (aOR 1.05 [95%CI:0.38,2.85]) or mean difference in hemoglobin concentration (g/dl) (0.04 [95%CI:-0,0.39) (Supplementary Table S3). However, baseline sub-patent infections persisted, with these infections increasing in density leading to detection by microscopy (aOR 9.54 [95%CI:5.42,16.8]) and PCR (aOR 8.03 [95%CI:5.50,11.7]) during follow-up. Thirty-four percent (20/59) of students with sub-patent infections at baseline had positive RDTs at the six-week follow-up, suggesting that repeated screening-and-treatment may further reduce prevalence of infection.

## Discussion

School-based screening-and-treatment is an attractive strategy to decrease the burden of malaria in school-age children. Our results show this strategy successfully decreased the burden of *Pf* infection, nearly halving the prevalence of infections detected by microscopy. However, the ability of RDTs to detect all infections varied by transmission setting, contributing to differences in the impact of the intervention. In lower, seasonal transmission settings, a large proportion of infections went untreated because they were sub-patent. While in higher transmission settings, the duration of the impact of screening-and-treatment was limited by new infections.

Overall, our results provide insights on which settings and for which outcomes screening-and-treatment warrants further consideration as an approach to deliver school-based preventive treatment. The only published clinical trial evaluating school-based screening-and-treatment showed no long-term impact on *Pf* infection, anemia, or cognition.^17^ However, the average transmission in the schools in that cluster-randomized trial in coastal Kenya was similar to transmission intensity of the lower, seasonal prevalence schools our study, where RDTs failed to detect a large proportion of infections. While these sub-patent infections were not associated with clinical disease in our six-week follow-up period, they likely contribute to the lack of long term impact. The lack of efficacy of screening-and-treatment for other target populations, including malaria in pregnancy^24^ and community-wide interventions^25,26^ has also been attributed to the low sensitivity of currently available RDTs. The sensitivity and specificity of RDTs in our study were similar the prior school-based screen-and-treat trial and other community-based studies.^17,27–29^

The different patterns of infection between treated and untreated students in both transmission settings have important implications for the design of school-based malaria control interventions. Our findings suggest that in settings with perennial, moderate-to-high transmission, the screen-and-treat approach using currently available screening tools may be effective in identifying and treating most infections but the duration of the impact is short-lived due to the high frequency of new infection. Thus, in these settings the design of the intervention could be improved by either repeating the intervention or by using a longer acting drug, such as dihydroartemisinin-piperaquine.

In lower transmission settings where a high proportion of infections fall below the level of detection of commonly used point-of-care tests, a screening test with higher sensitivity than a standard RDT should improve the effectiveness of the screen-and-treat approach. Newly approved point-of-care tests are currently undergoing evaluation and may be suitable for use in school-based interventions, particularly in low transmission settings.^30^

The alternative to screening-and-treatment is providing treatment to all students without screening, as in intermittent preventive treatment, an approach used widely to prevent the adverse effects of malaria during pregnancy and for seasonal malaria prevention in children in some settings. In recent meta-analyses of preventive treatment in schoolchildren, the majority of studies provided treatment to all students enrolled in the intervention arm and showed substantial reductions of both *Pf* infection prevalence and clinical malaria.^31^ This approach has the advantages of providing prophylaxis to all students regardless of their infection status and is easier to implement as no blood collection or time for testing is required. However, there are concerns about the cost-benefit of this approach if prevalence is low and the impact of selective pressure on drug resistant parasite resulting from such widespread administration of antimalarial drugs.

While our findings provide new insights into the utility of school-based screen-and-treat approaches, there are some limitations. We designed our observational study to investigate the dynamics of infections in treated and untreated children after screening. While randomized trials provide more definitive conclusions about the efficacy of the intervention, our approach nevertheless enabled assessment of the effect of the intervention on parasite prevalence in students who did and did not receive treatment based on RDT results in varied transmission settings. However, the absence of a control arm complicates interpretation of some results. In the overall study population, prevalence of anemia was significantly lower six weeks after the intervention. However, in analyses that stratified students by baseline anemia and treatment status, the intervention was not associated with lower anemia at six weeks. This finding is likely due to our inability to control for complex confounders (e.g. nutritional status), and secular trends leading to changes in anemia prevalence not related to the intervention. Furthermore, the study involved only six weeks of follow-up, limiting our ability to comment on the longer-term impacts of sub-patent infections.

Given these results, the utility of the screen-and-treat approach using currently available RDTs must be evaluated based on the goal of treatment. Sub-patent infections were not associated with clinical disease over the course of follow-up in this population, and the intervention reduced the prevalence of infections detected by microscopy, suggesting that screening-and-treatment may be sufficient to reduce clinical malaria. However, screening-and-treatment failed to reduce anemia and, therefore, may not reduce the impact of malaria on cognitive function and education. The impact preventive treatment of malaria on these endpoints may be increased by either targeting all students with antimalarial medication through intermittent preventive treatment, and/or combining malaria preventive treatment with other interventions, such as deworming and nutrition programs, to jointly tackle the multiple etiologies of anemia in school-age children.

While the primary aim of school-based malaria control is to decrease the adverse consequences of *Pf* infection in students, there may also be indirect effects on transmission in the community.^32,33^ Increasing evidence suggests both school-age children and persistent infections contribute significantly to malaria transmission.^34–36^ As such, studies of school-based malaria control interventions should also measure the indirect effects on malaria transmission in the community.

Given the high prevalence of *Pf* infection and the negative impacts of infection on the health and educational achievement of school-age children, development of strategies to decrease this burden is critical. School-based screening-and-treatment effectively reduced *Pf* infection in our study and is an attractive strategy as treatment is targeted to students with documented infection. Further studies are needed to evaluate this approach in different transmission settings as well as the utility of more sensitive screening tests for this purpose.

## Data Availability

After publication, data will be made available through publicly available databases.

## Acknowledgements

We thank Heidi Fancher and the administrative team at the Malaria Alert Center for their support in executing the study. We are grateful for the hard work and dedication of the field team, study nurses, teachers, school administrators who made the study possible. Most importantly, we thank the participants for their commitment and patience.

