## Supplemental Information for "Impact of school-based malaria screening and treatment on *P. falciparum* infection and anemia prevalence in two transmission settings in Malawi"

**Contents:**

Table S1: Log-binomial model of sub-patent malaria infection at baseline

Figure S1: Proportion of *P. falciparum* infections detected by microscopy (A) and PCR (B) overall and stratified by school

Table S2: Log-binomial model of anemia at baseline

Figure S2: Unadjusted prevalence of anemia six weeks after the intervention by baseline anemia and treatment

Table S3: The association of baseline sub-patent infections and outcomes during follow-up

Table S4: STROBE Checklist

| Table S1. Associations with sub-patent malaria infection at baseline^1^ | | | |
| --- | --- | --- | --- |
|  | **n(%)^2^** | **Unadjusted Prevalence Ratio** | **Adjusted Prevalence Ratio** |
| School |  |  |  |
| Bvumbwe | 12(7.1) | 1 | 1 |
| Ngowe | 10(7.7) | 1.0 (0.5, 2.3) | 1.1 (0.5, 2.5) |
| Maseya | 22(23.4) | 3.0 (1.6, 5.8) | 3.1 (1.6, 6.0) |
| Makhuwira | 19(35.9) | 4.7 (2.5, 9.0) | 4.9 (2.6, 9.3) |
| Season |  |  |  |
| Rainy | 26(12.7) | 1 | 1 |
| Dry | 37(15.4) | 1.3 (0.8, 2.0) | 1.2 (0.8, 1.8) |
| Age in years |  |  |  |
| 5 to 9 | 12(9.2) | 1 | 1 |
| 10 to 15 | 51(16.2) | 1.6 (0.9, 2.9) | 1.5 (0.9, 2.7) |
| Sex |  |  |  |
| Male | 25(13.8) | 1 | 1 |
| Female | 38(14.4) | 1.0 (0.7, 1.7) | 1.0 (0.7, 1.6) |
| Anemic^3^ |  |  |  |
| No | 54(13.4) | 1 |  |
| Yes | 9(21.9) | 1.7 (0.9, 3.1) |  |
| Fever last 48 hours^4^ |  |  |  |
| No | 44(13.4) | 1 |  |
| Yes | 19(17.6) | 1.3 (0.8, 2.2) |  |
| Recent malaria treatment^5^ |  |  |  |
| No | 55(14.4) | 1 |  |
| Yes | 8(13.6) | 0.9 (0.5, 1.8) |  |
| Slept under bed net last night |  |  |  |
| No | 35(14.3) | 1 |  |
| Yes | 28(14.0) | 0.9 (0.6, 1.5) |  |
| 1. Log-binomial model with school, season, age and sex retained in model *a priori,* all other variables retained in model if *p*<0.05; with the exception of missing data, univariate association were assessed in all children with a negative RDT at baseline (n=445), among whom 5 were missing data for recent medication, and 8 were missing data for fever in last 48 hours. The final model include 437 children.  2. Number and percent of children with sub-patent malaria infection at baseline result within each strata  3. Defined as hemoglobin less than 11.0 g/dL  4. Fever in the last 48 hours reported or measured temperature $\geq$37.5ºC  5. Recent treatment defined as student or parent reported receipt of an effective antimalarial drug in the two weeks preceding the interview | | | |

Figure S1: Proportion of *P. falciparum* infections detected by microscopy (A) and PCR (B) overall and stratified by school.

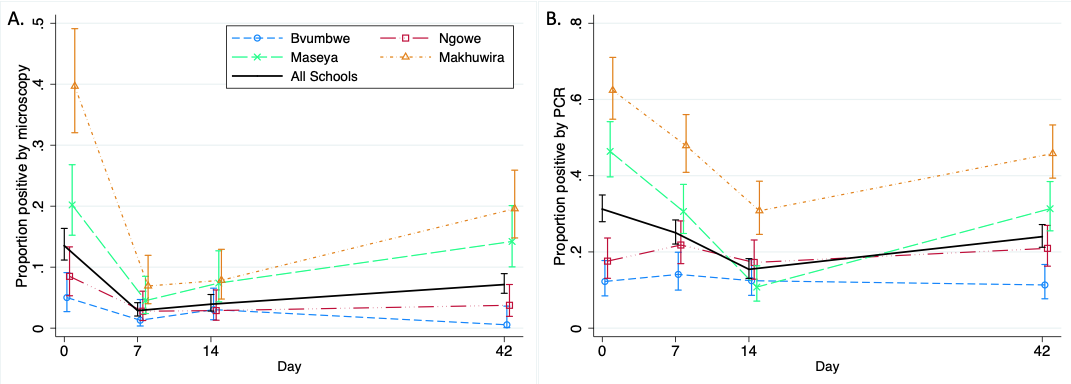

| Table S2: Associations with anemia at baseline^1^ | | | |
| --- | --- | --- | --- |
|  | **Prevalence n(%)^2^** | **Unadjusted Prevalence Ratio** | **Adjusted Prevalence Ratio** |
| School |  |  |  |
| Bvumbwe | 9(4.9) | 1 | 1 |
| Ngowe | 35(18.8) | 3.6 (1.8, 7.3) | 2.5(1.2, 5.2) |
| Maseya | 34(18.4) | 3.7 (1.8, 7.5) | 2.2(1.1, 4.7) |
| Makhuwira | 28(18.8) | 3.8 (1.9, 7.8) | 2.2(1.0, 4.8) |
| Season |  |  |  |
| Rainy | 61(16.8) | 1 | 1 |
| Dry | 45(13.2) | 0.8 (0.5, 1.1) | 0.9(0.6, 1.2) |
| Age |  |  |  |
| 5-9 years | 31(16.3) | 1 | 1 |
| 10-15 years | 75(14.6) | 0.9 (0.6, 1.3) | 0.8(0.6, 1.2) |
| Sex |  |  |  |
| Male | 63(19.6) | 1 | 1 |
| Female | 43(11.3) | 0.6 (0.4, 0.8) | 0.6(0.5, 0.9) |
| RDT at baseline |  |  |  |
| Negative | 41(9.2) | 1 | 1 |
| Positive | 65(25.2) | 2.7 (1.9, 3.9) | 2.1(1.4, 3.2) |
| Fever last 48 hours^3^ |  |  |  |
| No | 63(12.6) | 1 | 1 |
| Yes | 40(20.9) | 1.7(1.2, 2.4) | 1.5(1.0, 2.1) |
| Recent malaria treatment^4^ |  |  |  |
| No | 77(13.4) | 1 |  |
| Yes | 27(22.3) | 1.6(1.1, 2.5) |  |
| Slept under bed net last night |  |  |  |
| No | 60(15.2) | 1 |  |
| Yes | 46(14.9) | 0.9 (0.7, 1.4) |  |
| 1. Log-binomial model of anemia (defined as hemoglobin <11.0g/dL) with school, season, age and sex retained in model *a priori*, all other variables retained in model if *P*<.05. One child was missing data for anemia. With the exception of missing data, univariate associations were assessed in 703 children, among whom 9 were missing data for recent medication, and 13 were missing data for fever in last 48 hours. The final model included 690 children.  2. Number and percent of children with anemia at baseline result within each strata  3. Fever in the last 48 hours reported or measured temperature $\geq$37.5ºC  4. Recent treatment defined as student or parent reported receipt of an effective antimalarial drug in the two weeks preceding the interview | | | |

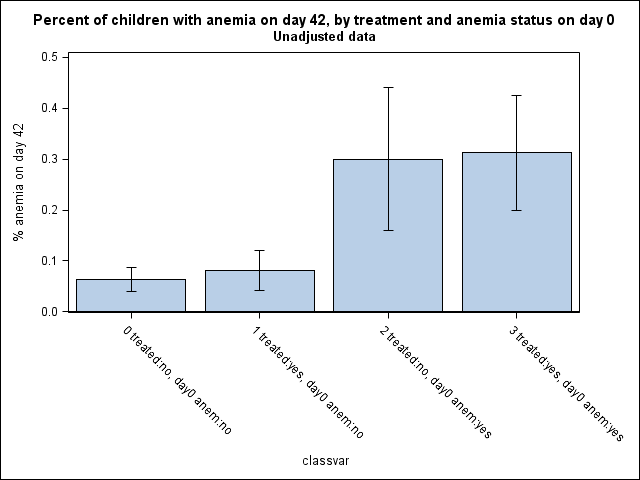

Figure S2: Unadjusted prevalence of anemia six weeks after the intervention by baseline anemia and treatment

Not treated	Treated	Not treated	Treated

| Not anemic at baseline | | Anemic at baseline. |

Prevalence of anemia post-intervention

| Table S3. The association of baseline sub-patent infections and outcomes during follow-up | | |
| --- | --- | --- |
|  | **Adjusted OR^1^ (95% CI)** | ***P* value** |
| Fever in the last 48 hours^2^ during follow-up | 1.05 (0.64, 1.72) | 0.8547 |
| Malaria treatment^3^ during follow-up | 0.97 (0.40, 2.32) | 0.9383 |
| Infection detected by microscopy during follow-up | 9.5 (5.4, 16.8) | <.0001 |
| Infection detected by PCR during follow-up | 8.0 (5.5, 11.7) | <.0001 |
| Anemic^4^ at day 42 | 1.05 (0.38, 2.85) | 0.9287 |
|  | **Beta-coeff (95% CI)** |  |
| Hemoglobin at day 42 | 0.04 (-0.31, 0.39) | 0.8164 |
| 1. Models assessed outcomes for 445 children with a negative RDT test at baseline. Logistic GEE models with an unstructured covariance for children nested in schools were used to model reported fever, microscopy-detected malaria, and PCR-detected malaria. A simple logistic model was used to model anemia at day 42, and a linear model was used to model hemoglobin at day 42. All models adjusted *a priori* for school, season, age less than 10 years versus 10 or older, and anemia at baseline.  2. Fever in the last 48 hours reported or measured temperature $\geq$37.5ºC  3. Recent treatment defined as student or parent reported receipt of an effective antimalarial drug in the two weeks preceding the interview at each visit  4. Defined as hemoglobin less than 11.0 g/dL | | |

| Table S4: STROBE Checklist | | |  |
| --- | --- | --- | --- |
| **Section/topic** | Item No | Checklist item | Reported on page # |
| **Title and abstract** | 1 | (*a*) Indicate the study’s design with a commonly used term in the title or the abstract | 1 |
|  |  | (*b*) Provide in the abstract an informative and balanced summary of what was done and what was found | 2 |
| Introduction | | |  |
| Background/rationale | 2 | Explain the scientific background and rationale for the investigation being reported | 3-4 |
| Objectives | 3 | State specific objectives, including any prespecified hypotheses | 4 |
| Methods | | |  |
| Study design | 4 | Present key elements of study design early in the paper | 4-6 |
| Setting | 5 | Describe the setting, locations, and relevant dates, including periods of recruitment, exposure, follow-up, and data collection | 5 |
| Participants | 6 | (*a*) Give the eligibility criteria, and the sources and methods of selection of participants. Describe methods of follow-up | 5 |
|  |  | (*b*) For matched studies, give matching criteria and number of exposed and unexposed |  |
| Variables | 7 | Clearly define all outcomes, exposures, predictors, potential confounders, and effect modifiers. Give diagnostic criteria, if applicable | 6-7 |
| Data sources/ measurement | 8* | For each variable of interest, give sources of data and details of methods of assessment (measurement). Describe comparability of assessment methods if there is more than one group | 5-6 |
| Bias | 9 | Describe any efforts to address potential sources of bias | 5 |
| Study size | 10 | Explain how the study size was arrived at | 6 |
| Quantitative variables | 11 | Explain how quantitative variables were handled in the analyses. If applicable, describe which groupings were chosen and why | 7 |
| Statistical methods | 12 | (*a*) Describe all statistical methods, including those used to control for confounding | 7 |
|  |  | (*b*) Describe any methods used to examine subgroups and interactions | 7-8 |
|  |  | (*c*) Explain how missing data were addressed |  |
|  |  | (*d*) If applicable, explain how loss to follow-up was addressed |  |
|  |  | (*e*) Describe any sensitivity analyses |  |
| Results | | |  |
| Participants | 13* | (a) Report numbers of individuals at each stage of study—eg numbers potentially eligible, examined for eligibility, confirmed eligible, included in the study, completing follow-up, and analysed | 8, Figure 1 |
|  |  | (b) Give reasons for non-participation at each stage | Figure 1 |
|  |  | (c) Consider use of a flow diagram | Figure 1 |
| Descriptive data | 14* | (a) Give characteristics of study participants (eg demographic, clinical, social) and information on exposures and potential confounders | Table 1 |
|  |  | (b) Indicate number of participants with missing data for each variable of interest | Table 1 |
|  |  | (c) Summarise follow-up time (eg, average and total amount) | 8, Figure 1 |
| Outcome data | 15* | Report numbers of outcome events or summary measures over time | 8-9, Figures 2, S1 |
| Main results | 16 | (*a*) Give unadjusted estimates and, if applicable, confounder-adjusted estimates and their precision (eg, 95% confidence interval). Make clear which confounders were adjusted for and why they were included | Tables 2, 3, S1, S2, S3 |
|  |  | (*b*) Report category boundaries when continuous variables were categorized | NA |
|  |  | (*c*) If relevant, consider translating estimates of relative risk into absolute risk for a meaningful time period |  |
| Other analyses | 17 | Report other analyses done—eg analyses of subgroups and interactions, and sensitivity analyses |  |
| Discussion | | |  |
| Key results | 18 | Summarise key results with reference to study objectives | 11 |
| Limitations | 19 | Discuss limitations of the study, taking into account sources of potential bias or imprecision. Discuss both direction and magnitude of any potential bias | 13 |
| Interpretation | 20 | Give a cautious overall interpretation of results considering objectives, limitations, multiplicity of analyses, results from similar studies, and other relevant evidence | 15 |
| Generalisability | 21 | Discuss the generalisability (external validity) of the study results | 14 |
| Other information | | |  |
| Funding | 22 | Give the source of funding and the role of the funders for the present study and, if applicable, for the original study on which the present article is based | 15 |
